# Antiemetic effect of pregabalin in breast reconstruction surgery in patients after bariatric surgery: Prospective, randomized, double-blind study

**DOI:** 10.1101/2023.07.20.23292941

**Authors:** Rafael Reis Fernandes, Marcello Fonseca Salgado-Filho, Guilherme Bracco Graziosi, André Luiz Viana Nery da Silva, Flávio Augusto Amaral Fernandes Távora, Caio Pontes de Azevedo, Alice Ramos Oliveira da Silva, Nubia Verçosa, Ismar Lima Cavalcanti

## Abstract

**Introduction:** Postoperative nausea and vomiting (PONV) is a common complication after general anesthesia. Pregabalin may reduce its incidence. The objective of this study was to evaluate the adjuvant antiemetic effect of pregabalin in the first 24 hours after surgery in patients undergoing breast reconstruction after bariatric surgery.

**Materials and Methods:** This prospective, randomized, double-blind study had 52 female patients aged between 18 and 64 years with physical status 1-2 of the American Society of Anesthesiologists who underwent breast reconstruction after bariatric surgery. The patients were divided into two groups. Patients in the control group received placebo, and those in the pregabalin group received 75 mg of pregabalin 2 hours before surgery and 75 mg 12 hours later. All patients received 4 mg dexamethasone and 4 mg ondansetron. The incidence of PONV was evaluated in the immediate postoperative period and 6 hours, 12 hours, and 24 hours later. The need for rescue doses of antiemetics and adverse events in the first 24 hours were recorded.

**Results:** The groups were homogeneous in clinical and treatment variables. There was no significant difference in the incidence of PONV over time in the control group (P = 0.71/no occurrence) or in the pregabalin group (P = 0.11/P = 0.26). There was no significant difference in the need for rescue antiemetic dose (P = 0.40) or in the incidence of adverse events (P = 0.51) between groups.

**Conclusion:** The administration of pregabalin in the first 24 hours after surgery did not significantly reduce PONV in patients undergoing breast reconstruction after bariatric surgery.

## Introduction

Postoperative nausea and vomiting (PONV) is common after general anesthesia, with an incidence ranging from 30 to 50% [1]. Thus, prevention measures are routinely taken during anesthetic-surgical procedures, especially those performed under general anesthesia using opioids, inhalational anesthetics, or nitrous oxide [1,2]. The goals of preventive treatment for PONV are to avoid unexpected hospitalizations, delays in hospital discharge, increases in hospital costs, and patient discomfort and dissatisfaction [3,4]. Therefore, an aggressive and multimodal prophylactic approach is recommended, especially in high-risk patients. These high-risk patients are classified using the simplified Apfel score [2,5,6]. Recently, GABAergic adjuvants such as pregabalin have been associated with multimodal management for better control of PONV [1,7].

We hypothesized that patients who were preventively medicated with pregabalin would have a lower rate of nausea and vomiting in the first 24 postoperative hours. The objective of this study was to evaluate the antiemetic effect of pregabalin in the first 24 hours after surgery in patients undergoing breast reconstruction after bariatric surgery under general anesthesia.

## Materials and Methods

After approval by the Ethics Committee in Human Research of the Antônio Pedro University Hospital, Niterói, Rio de Janeiro, Brazil, on November 13, 2018 (CAAE: 85415718.2.0000.5243) and registration in the Brazilian Clinical Trials Registry (ReBEC – UTN: U1111-1259-6153), a prospective, randomized, double-blind, placebo-controlled, single-center clinical trial with 52 adult patients undergoing elective breast reconstruction after bariatric surgery under general anesthesia in a tertiary hospital between August 2019 and July 2021 was conducted. All patients signed a written informed consent form before surgery. This manuscript follows the guidelines of CONSORT.

The inclusion criteria were 52 female patients aged between 18 and 64 years, with physical status classified by the American Society of Anesthesiologists (ASA) scale 1 and 2 [8], who underwent breast reconstruction after bariatric surgery. The exclusion criteria were patients who had participated in another study in the last month; smokers; alcohol drinkers; body mass index greater than 35 mg/m^2^; occurrence of episodes of nausea or vomiting in the 24 hours before surgery; use of corticosteroids; use of psychoactive drugs or any drug with an antiemetic effect; known hypersensitivity to any study medication; patients with severe kidney, liver, lung, heart, brain, or bone marrow diseases; and patients who had a history of uncontrollable vomiting even after administration of rescue medication.

No patient received any premedication. In the operating room, the patients were monitored with pulse oximetry, noninvasive blood pressure, electrocardioscopy, capnography, and degree of neuromuscular blockade. A 20-G intravenous cannula was inserted into the right or left arm. Preoxygenation was performed with 100% oxygen for 5 minutes at 6 L·min^-1^ using a face mask. Both groups underwent the same general anesthesia. Anesthesia was induced with intravenous administration of 1.5 mg.kg^-1^ fentanyl (Cristália^®^, São Paulo, Brazil) 3 mcg·kg^-1^, lidocaine (Cristália^®^, São Paulo, Brazil), 1-2 mg·kg^-1^ propofol (Cristália^®^, São Paulo, Brazil), and 0.6 mg·kg^-1^ rocuronium bromide (Cristália^®^, São Paulo, Brazil) followed by orotracheal intubation. Anesthesia was maintained with inhalation of sevoflurane (Cristália^®^, São Paulo, Brazil) at a 2% end-tidal concentration combined with 50% oxygen/compressed air at 1 L·min^-1^ and 0.05 to 0.3 mcg·kg^-1^·min^-1^ remifentanil (GSK^®^, Rio de Janeiro, Brazil) intravenously by an infusion pump (B. Braun^®^, Rio de Janeiro, Brazil) when necessary. The patients received perioperative preventive analgesia with the nonsteroidal anti-inflammatory drug parecoxib sodium (Pfizer^®^, São Paulo, Brazil) 40 mg intravenously and dipyrone sodium (Sanofi Aventis^®^, São Paulo, Brazil) 50 mg·kg^-1^ intravenously. For perioperative preventive antiemesis, the patients received 4 mg intravenous ondansetron (Cristália^®^, São Paulo, Brazil) and 4 mg intravenous dexamethasone (Aché^®^, São Paulo, Brazil). At the end of surgery, 2 mg·kg^-1^ of sugammadex (MSD^®^, São Paulo, Brazil) was administered intravenously to reverse the neuromuscular block, and the trachea was extubated. Postoperative analgesia was administered with 50 mg·kg^-1^ intravenous dipyrone (Sanofi Aventis^®^, São Paulo, Brazil) every 4 hours and 40 mg intravenous parecoxib sodium (Pfizer^®^, São Paulo, Brazil) every 12 hours in regular doses and 10 mg intravenous nalbuphine hydrochloride (Cristália^®^, São Paulo, Brazil) as a rescue dose every 4 hours when the visual analog scale (VAS) was greater than 4 (moderate pain) or when requested by the patient due to uncontrolled pain. Postoperative antiemesis was performed with rescue doses of 4 mg ondansetron (Cristália^®^, São Paulo, Brazil) every 8 hours, as requested by the patient and after nausea or vomiting.

The primary outcome of the study was the antiemetic effect of pregabalin. The frequency and intensity of individual episodes of nausea and the frequency of vomiting immediately, 6 hours, 12 hours, and 24 hours after surgery were compared. The secondary outcomes of the study were the rating of pain by VAS at rest and on movement at the same four times, the amounts of antiemetic and analgesic rescue medications used in the first 24 postoperative hours, the need for opioid medication after hospital discharge, chronic pain 3 months after surgery [9], and the incidence of adverse events in the first 24 postoperative hours.

For the purpose of this study, nausea was defined as a subjective unpleasant and involuntary sensation of the urge to vomit, without the expulsion of stomach contents, and vomiting was defined as the expulsion of stomach contents through the mouth [1].

Patients from both groups were evaluated at four times during the first 24 hours postoperatively, immediately, 6 hours, 12 hours, and 24 hours, to record PONV onset, pain at rest or movement according to the VAS, the need for antiemetic and/or analgesic rescue medication, and the occurrence of adverse events. The rescue doses of nalbuphine in the first 24 hours, the consumption of remifentanil during the perioperative period, and the need for opioid use after hospital discharge were compared between the groups to evaluate the opioid-sparing effect of pregabalin. Three months after hospital discharge, all patients were contacted by telephone to evaluate postoperative chronic pain.

A protocol form was used to record the data. It contained the patient’s medical record number, initials, age, anthropometric data, previous pathological history, physical status according to the ASA [8], total procedure time, total anesthesia time, total waking time (which was the time between interruption of anesthesia and extubation), amount of fluids administered, and the Aldrete and Kroulik scale [10] of postanesthetic recovery. The frequency of PONV at each time, as well as pain, according to the VAS, during the rest period and during movement after surgery were also recorded in the protocol form. The antiemetic and analgesic rescue medications used in the first 24 postoperative hours, the opioid sparing effect through quantification of the rescue doses of nalbuphine hydrochloride in the first 24 hours, the consumption of remifentanil in the perioperative period and the need for opioids after hospital discharge, the evaluation of the development of chronic pain after 3 months postoperatively, and the occurrence of adverse events in the first 24 hours were recorded.

### Statistical analysis

To calculate the sample size and power of the study, Grant et al. [1], who performed a meta-analysis on pregabalin reducing PONV, obtained a 53% reduction in the incidence of PONV in the group that used pregabalin. Thus, to calculate a 53% reduction in PONV cases with a study power of 90%, with an alpha value less than 0.05 and a beta value less than 0.2, 24 patients were required in each group. An additional 10% was included to cover possible case losses, for a total of 52 patients. GraphPad Prism QuickCalcs^®^ software (Inc., La Jolla, CA, USA) was used for this calculation.

After electronic randomization (Randomized ^®^), the patients were divided into two groups of 26 in a 1:1 ratio: the control group and the pregabalin group. The compounding pharmacy of the hospital formulated a placebo capsule consisting of starch and another capsule of 75 mg of pregabalin. The pharmacist allocated the tablets (placebo or pregabalin) into envelopes of identical appearance, numbered from 1 to 52, following randomization principles. Each envelope contained two placebo tablets or two pregabalin tablets. One tablet was given to the patient 2 hours before surgery in the preoperative period and one tablet 12 hours later. The researcher and the patient were blinded to the pills administered.

For descriptive analysis, we organized the observed data into tables, and they are expressed as measures of central tendency and dispersion suitable for numerical data and as frequency and percentage for categorical data.

The inferential analysis consisted of the following methods: Clinical and treatment variables were compared between groups (placebo and pregabalin) using the independent-sample Student’s t test (parametric) or the Mann‒Whitney test (nonparametric) for numerical data, and the χ^2^ test or Fisher’s exact test for categorical data. Repeated-measures analysis of variance (ANOVA) was run on categorical data using the CATMOD procedure [11] of SAS^®^ statistical software to verify whether there was significant variation in nausea or vomiting over the four time points. The longitudinal analysis of VAS over the four time points was composed of repeated-measures ANOVA within each group. Repeated-measures ANOVA with one factor, highlighting the interaction component (time × group), was done to compare the evolution of VAS between the groups. The Bonferroni test of multiple comparisons was applied to identify which times differed significantly from each other within each group.

The Shapiro‒Wilk test was run to see if each variable conformed to the normal distribution, together with the graphical analysis of the histograms. The level of significance adopted was 5%. Statistical analysis was performed with SPSS statistical software version 26 and SAS^®^ 6.11 (SAS Institute, Inc., Cary, NC).

## Results

A total of 52 eligible female patients were recruited and randomized into two groups: 26 to the control group and 26 to the pregabalin group (Fig. 1). All included patients were allocated to a group. There was no loss in the analyses, and there was no loss to follow-up.

**Fig. 1.**
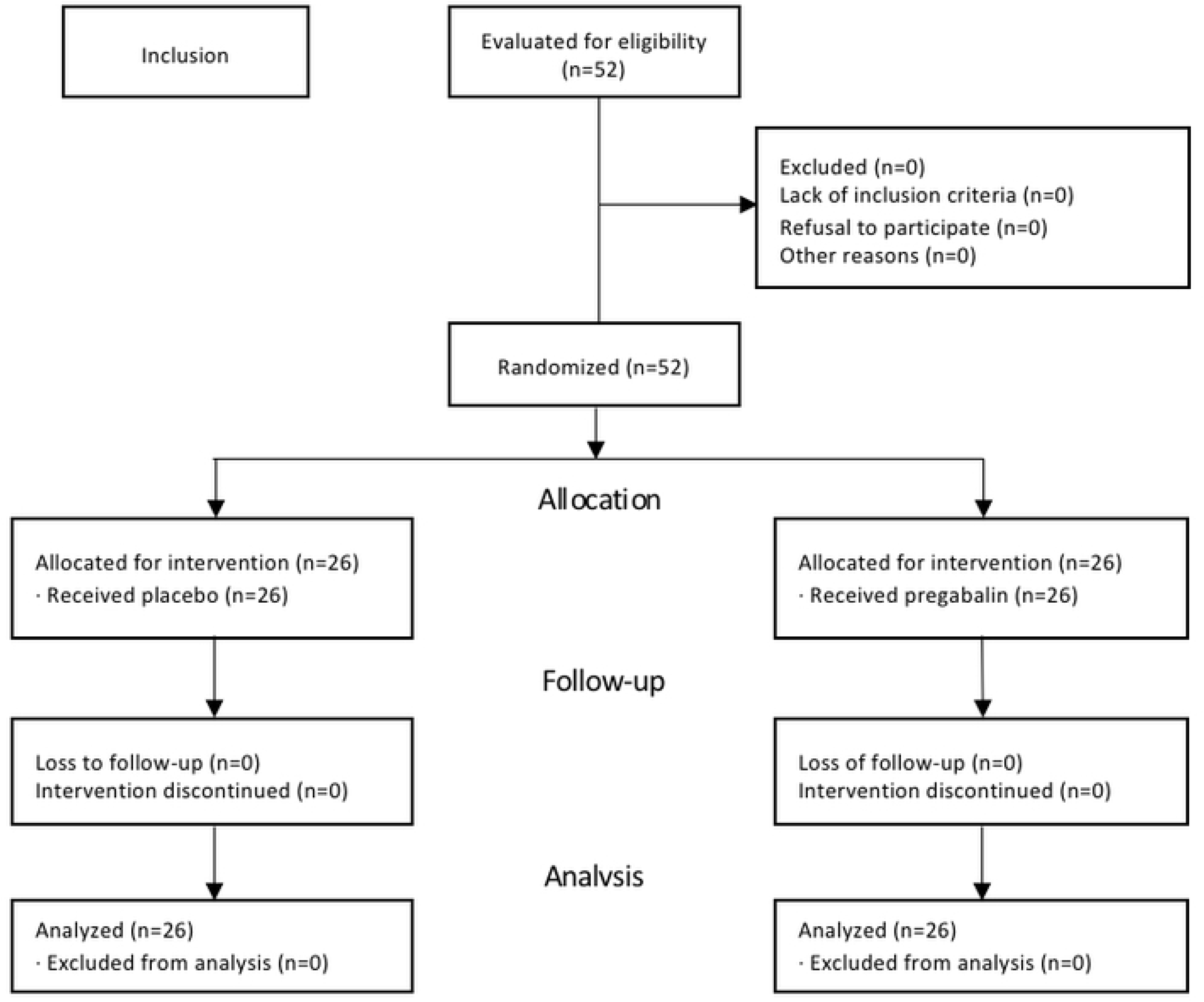
Study patient flowchart - CONSORT.

There was no significant difference at the 5% level between the groups in the clinical variables or the characteristics of surgery between the groups. Demographic data were homogeneous, except for height, but this had no clinical significance (Table 1). There was no significant difference in the use of opioids in the perioperative period.

**Table 1.**
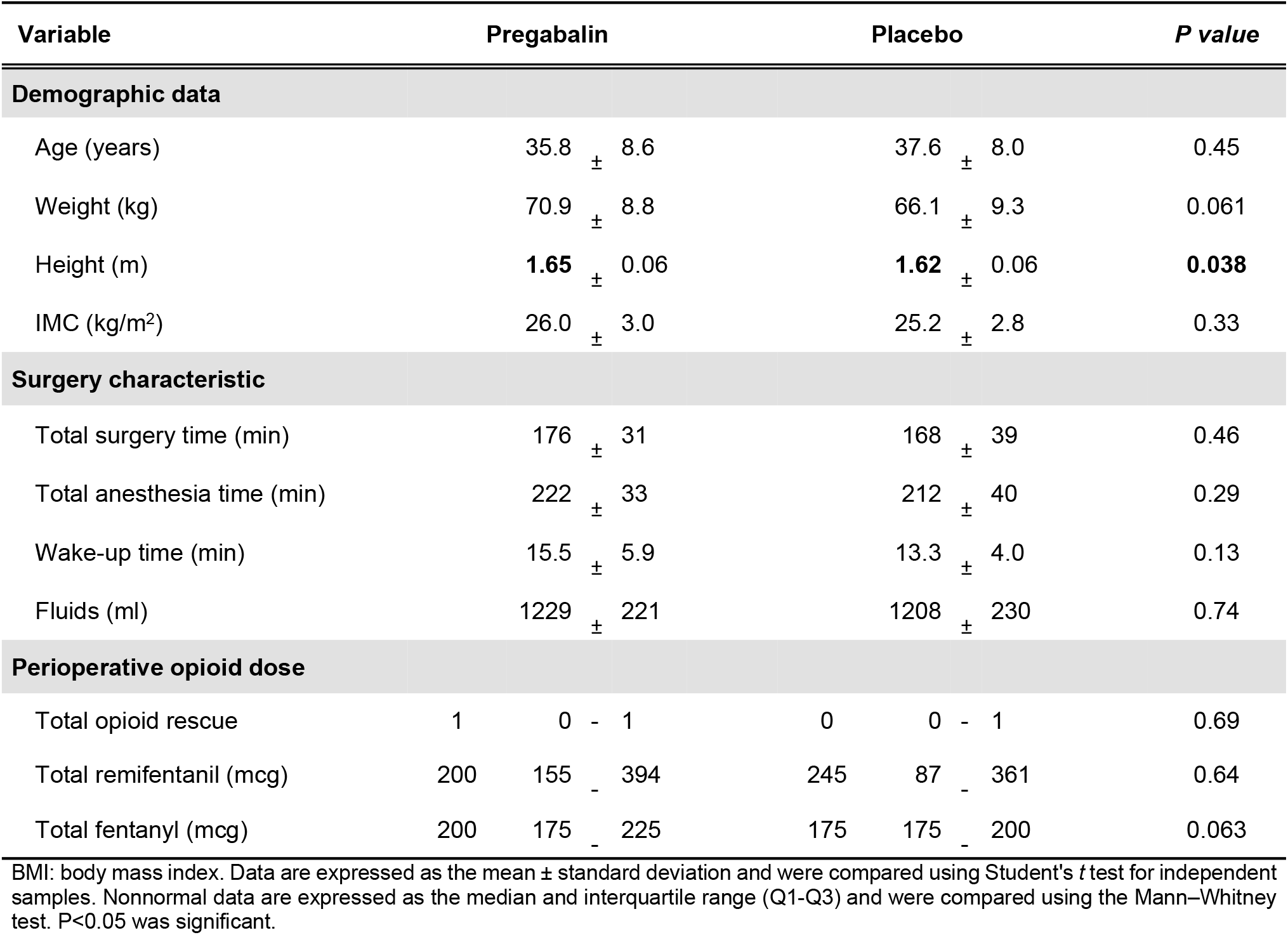
Demographic data, surgery characteristics and perioperative use of opioids.

| Variable | Pregabalin |  | Placebo |  | P value |
| --- | --- | --- | --- | --- | --- |
| Demographic data |  |  |  |  |  |
| Age (years) | 35.8 | ± 8.6 | 37.6 | ± 8.0 | 0.45 |
| Weight (kg) | 70.9 | ± 8.8 | 66.1 | ± 9.3 | 0.061 |
| Height (m) | 1.65 | ± 0.06 | 1.62 | ± 0.06 | 0.038 |
| IMC (kg/m²) | 26.0 | ± 3.0 | 25.2 | ± 2.8 | 0.33 |
| Surgery characteristic |  |  |  |  |  |
| Total surgery time (min) | 176 | ± 31 | 168 | ± 39 | 0.46 |
| Total anesthesia time (min) | 222 | ± 33 | 212 | ± 40 | 0.29 |
| Wake-up time (min) | 15.5 | ± 5.9 | 13.3 | ± 4.0 | 0.13 |
| Fluids (ml) | 1229 | ± 221 | 1208 | ± 230 | 0.74 |
| Perioperative opioid dose |  |  |  |  |  |
| Total opioid rescue | 1 | 0 - 1 | 0 | 0 - 1 | 0.69 |
| Total remifentanyl (mcg) | 200 | 155 - 394 | 245 | 87 - 361 | 0.64 |
| Total fentanyl (mcg) | 200 | 175 - 225 | 175 | 175 - 200 | 0.063 |

BMI: body mass index. Data are expressed as the mean ± standard deviation and were compared using Student's *t* test for independent samples. Nonnormal data are expressed as the median and interquartile range (Q1-Q3) and were compared using the Mann-Whitney test. *P*<0.05 was significant.

There was no significant difference between the groups in the categorical clinical variables, such as associated comorbidities, previous pathological history of pain, history of PONV, physical status classification by ASA, degree of consciousness in the preoperative period, operation time, heart rate at admission to the operating room, and Aldrete and Kroulik score at departure from the operating room (Table 2).

**Table 2.** Clinical categorical variables.

| Variable | Pregabalin |  | Placebo |  | P value |
| --- | --- | --- | --- | --- | --- |
|  | n | % | n | % |  |
| Comorbidities |  |  |  |  |  |
| yes | 9 | 34.6 | 9 | 34.6 | 1 |
| no | 17 | 65.4 | 17 | 65.4 |  |
| PPH Pain |  |  |  |  |  |
| yes | 0 | 0 | 0 | 0 | NA |
| no | 26 | 100 | 26 | 100 |  |
| PONV |  |  |  |  |  |
| yes | 1 | 3.8 | 4 | 15.4 | 0.17 |
| no | 25 | 96.2 | 22 | 84.6 |  |
| WING |  |  |  |  |  |
| I | 17 | 65.4 | 17 | 65.4 | 1 |
| II | 9 | 34.6 | 9 | 34.6 |  |
| Preop awareness |  |  |  |  |  |
| sleepy | 1 | 3.8 | 1 | 3.8 | 0.99 |
| calm | 18 | 69.2 | 17 | 65.4 |  |
| anxious | 7 | 26.9 | 8 | 30.8 |  |
| HR OR entry |  |  |  |  |  |
| < 60 bpm | 2 | 7.7 | 6 | 23.1 | 0.36 |
| 60-90 bpm | 17 | 65.4 | 15 | 57.7 |  |
| > 90 bpm | 7 | 26.9 | 5 | 19.2 |  |
| ALDRETE OR exit |  |  |  |  |  |
| 9 | 22 | 84.6 | 17 | 65.4 | 0.11 |
| 10 | 4 | 15.4 | 9 | 34.6 |  |

PPH: previous pathological history; PONV: postoperative nausea and vomiting; ASA: American Society of Anesthesiologists; Preoperative: preoperative period; HR: heart rate; OR: operating room. P<0.05 was significant ( $\chi^2$ test or Fisher's exact test).

There was no significant difference in the frequency of PONV between the groups at any time (Table 3).

**Table 3.** PONV episodes at the four times.

| Variable | Pregabalin |  | Placebo |  | *P value*<sup>2</sup> |
| --- | --- | --- | --- | --- | --- |
|  | n | % | n | % |  |
| Nausea |  |  |  |  |  |
| 0 h | 4 | 15.4 | 4 | 15.4 | 1 |
| 6 h | 5 | 19.2 | 4 | 15.4 | 0,50 |
| 12 h | 1 | 3.8 | 3 | 11.5 | 0,30 |
| 24 h | 6 | 23.1 | 6 | 23.1 | 1 |
| ANOVA repeated measures | *P value*<sup>1</sup> = 0.11 |  | *P value*<sup>1</sup> = 0.71 |  |  |
| Vomiting |  |  |  |  |  |
| 0 h | 2 | 7.7 | 0 | 0 | 0,25 |
| 6 h | 0 | 0 | 0 | 0 | NA |
| 12 h | 0 | 0 | 0 | 0 | NA |
| 24 h | 2 | 7.7 | 0 | 0 | 0,25 |
| ANOVA repeated measures | *P value*<sup>1</sup> = 0.26 |  | NA |  |  |

T: time; 0 h: immediate postoperative period; h: hours.
<sup>1</sup> ANOVA for repeated measures of categorical data by the CATMOD procedure of SAS® software.
<sup>2</sup> $\chi^2$ test or Fisher's exact test. NA: not applicable. $P < 0.05$ was significant.

There was no significant difference in VAS at rest or upon movement, at any time between the groups (Table 4). Within both groups, both at rest and in movement, there was a significant variation in pain control in the first 24 hours (Figs. 2 and 3).

**Figure 2.**
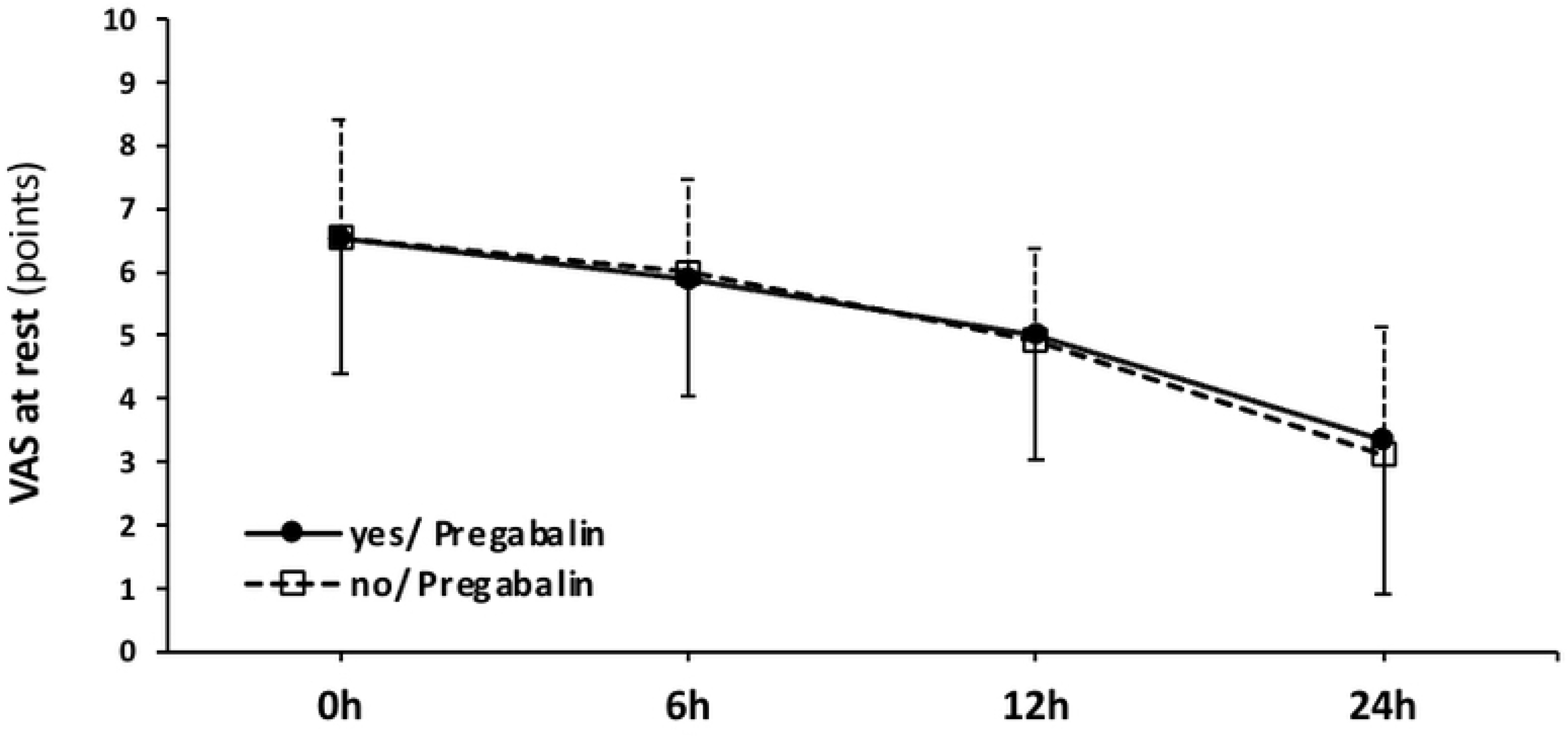
VAS at rest over time according to group.

**Figure 3.**
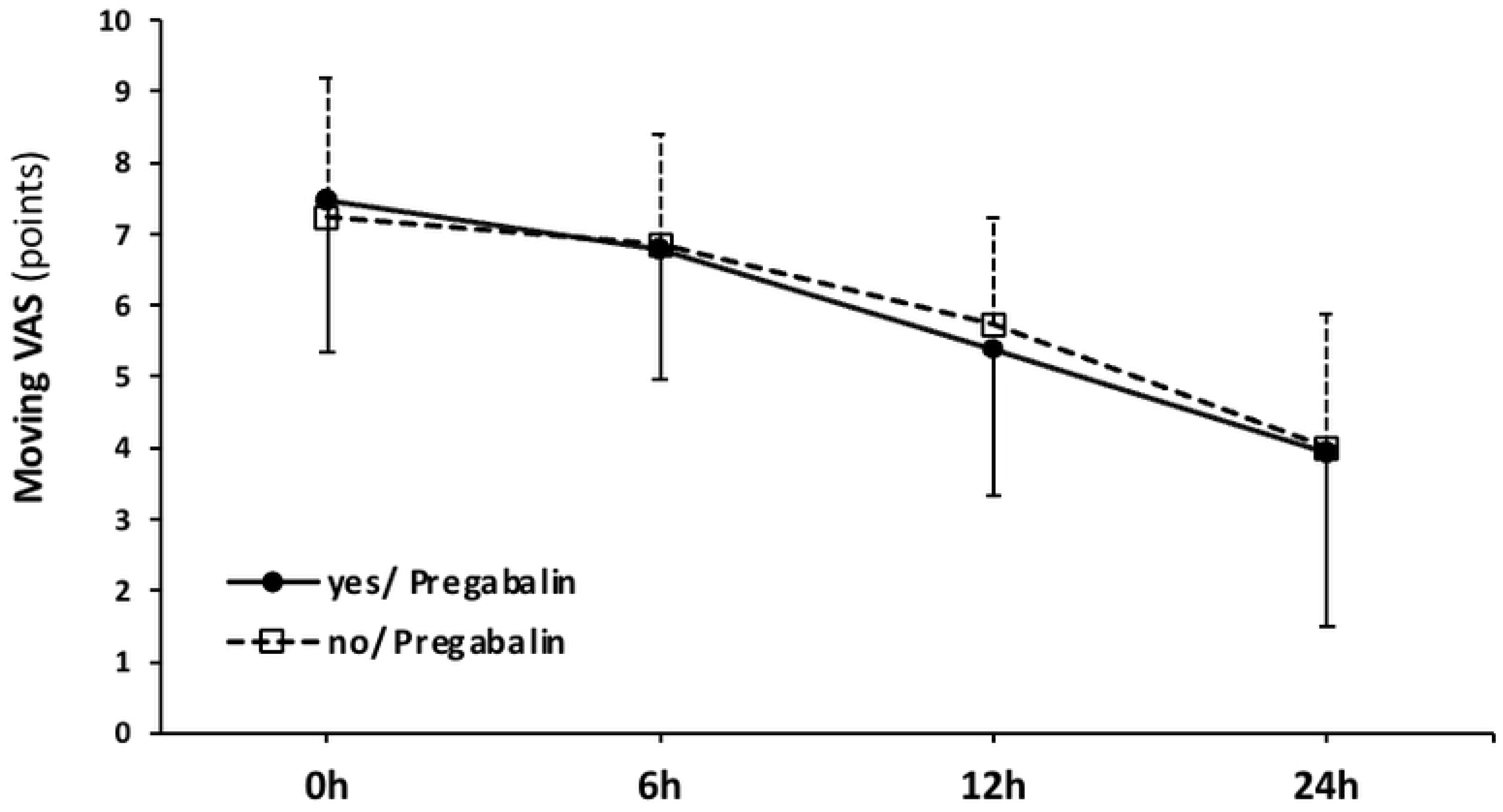
VAS in motion over time by group.

**Table 4.** Evolution of VAS at rest and during movement at the four times.

| Variable | Pregabalin | Placebo | *P value*<sup>2</sup> |
| --- | --- | --- | --- |
| **VAS at rest (points)** |  |  |  |
| 0 h | 6.5 ± 2.1 | 6.5 ± 1.9 | 0.99 |
| 6 h | 5.9 ± 1.8 | 6.0 ± 1.5 | 0.80 |
| 12 h | 5.0 ± 2.0 | 4.9 ± 1.4 | 0.87 |
| 24 h | 3.3 ± 2.4 | 3.1 ± 2.0 | 0.71 |
| *ANOVA repeated measures* | ***P value*<sup>1</sup> < 0.0001** | ***P value*<sup>1</sup> < 0.0001** |  |
| *Bonferroni multiple comparisons* | (0 h, 6 h) ≠ (12 h, 24 h)<br>and 12 h ≠ 24 h | (0 h, 6 h) ≠ (12 h, 24 h)<br>and 12 h ≠ 24 h |  |
| **Moving VAS (points)** |  |  |  |
| 0 h | 7.5 ± 2.1 | 7.2 ± 2.0 | 0.69 |
| 6 h | 6.8 ± 1.8 | 6.8 ± 1.5 | 0.87 |
| 12 h | 5.4 ± 2.1 | 5.7 ± 1.5 | 0.49 |
| 24 h | 3.9 ± 2.4 | 4.0 ± 1.9 | 0.90 |
| *ANOVA repeated measures* | ***P value*<sup>1</sup> < 0.0001** | ***P value*<sup>1</sup> < 0.0001** |  |
| *Bonferroni multiple comparisons* | (0 h, 6 h) ≠ (12 h, 24 h)<br>and 12 h ≠ 24 h | (0 h, 6 h) ≠ (12 h, 24 h)<br>and 12 h ≠ 24 h |  |

VAS: visual analog scale; 0 h: immediate postoperative period; h: hours.
<sup>1</sup> ANOVA for repeated measures within each group, time component. Bonferroni multiple comparisons test at the 5% level.
<sup>2</sup> Student's *t* test for independent samples.

There was no significant difference in the following variables between groups: frequency of adverse events in the first 24 hours after surgery, use of opioids after hospital discharge, use of opioid rescue medication or PONV in the first 24 hours after surgery, or development of postoperative chronic pain (Table 5).

**Table 5.** Postoperative variables by group.

| Variable | Pregabalin |  | Placebo |  | P value |
| --- | --- | --- | --- | --- | --- |
|  | n | % | n | % |  |
| Adverse events |  |  |  |  |  |
| yes | 5 | 19.2 | 7 | 26.9 | 0.51 |
| no | 21 | 80.8 | 19 | 73.1 |  |
| Postdischarge opioid use |  |  |  |  |  |
| yes | 9 | 37.5 | 8 | 36.4 | 0.94 |
| no | 15 | 62.5 | 14 | 63.6 |  |
| Total opioid rescue |  |  |  |  |  |
| none | 12 | 46.2 | 14 | 53.8 | 0.74 |
| one | 11 | 42.3 | 8 | 30.8 |  |
| 2 or 3 | 3 | 11.5 | 4 | 15.4 |  |
| PONV rescue |  |  |  |  |  |
| yes | 9 | 34.6 | 12 | 46.2 | 0.40 |
| no | 17 | 65.4 | 14 | 53.8 |  |
| Opioid rescue |  |  |  |  |  |
| yes | 13 | 50.0 | 11 | 42.3 | 0.58 |
| no | 13 | 50.0 | 15 | 57.7 |  |
| Chronic pain PO |  |  |  |  |  |
| yes | 6 | 25.0 | 9 | 40.9 | 0.25 |
| no | 18 | 75.0 | 13 | 59.1 |  |

PONV: postoperative nausea and vomiting; PO: postoperatively. P<0.05 was significant ( $\chi^2$ test or Fisher's exact test).

## Discussion

According to the Apfel criteria [6], the incidence of PONV in this study is moderate to high, ranging from 40 to 80%, since only female patients and nonsmokers were included in this study, factors that indicate aggressive multimodal treatment for the control of PONV symptoms [2,12]. In addition, these patients received general anesthesia with volatile anesthetics and opioids, which also increased the risk of PONV [12]. Adequate PONV control aims to avoid negative postoperative repercussions such as fluid and electrolyte imbalance, wound dehiscence, pulmonary aspiration, and prolonged hospital stay [4]. For this reason, advanced perioperative recovery protocols have been increasingly introduced into anesthetic practice to promote early recovery of patients by reducing the use of opioids, which in turn reduces PONV [1]. In the present study, we administered pregabalin adjuvantly because there is evidence that this drug reduces PONV [1,7]. Its mechanism of action in the prevention of PONV is not fully known, but it does block voltage-gated calcium channels by reducing the release of excitatory neurotransmitters that reduce postoperative inflammation. It has inhibitory action in the area postrema, which may help to reduce intraoperative and postoperative opioid consumption [1].

Pregabalin doses, dosages for use, and types of surgeries it is used for are heterogeneous [7]. This may explain the conflicting results of its use and suggests the need for more studies on the subject [7]. Pregabalin doses have ranged from 50 mg to 600 mg per day, with different dosages [1]. Some patients have used a dose of pregabalin preoperatively or postoperatively or a dose administered preoperatively and another 12 hours later [7,13]. Patients who have received higher doses of pregabalin have had less PONV, greater opioid-sparing effects, and greater efficacy of pain control; however, they have had more side effects [7,13]. In contrast, Mishriky et al. [14] found that a single preoperative dose of pregabalin was as effective as multiple doses and that low doses (less than 75 mg) were as effective as high doses (300 mg) in terms of reducing the consumption of morphine. For our study, we used 150 mg of pregabalin in the first 24 hours, divided into a 75 mg tablet 2 hours before surgery and a 75 mg tablet 12 hours after the first dose [15,16,17].

The total incidence of postoperative nausea in our study was 63.46%, all reported as mild, and the total incidence of postoperative vomiting was 7.69%. Previous studies [18, 19] reported that 5-HT3 receptor antagonists may be better antiemetics than anti-nausea agents, which may explain the much lower incidence of vomiting (7.69%) than the incidence of nausea (63,46%) in our study. At the four times studied (0 h, 6 hours, 12 hours, and 24 hours), there was no significant difference in PONV with the use of pregabalin. The total incidence of the use of rescue antiemetic medication in the first 24 hours after surgery was 40.38%, being slightly higher in the group that did not use pregabalin (P = 0.40). In their meta-analysis, Grant et al. [1] demonstrated that pregabalin reduces the incidence of nausea and vomiting and the administration of rescue antiemetics within the first 24 hours after surgery performed under general anesthesia. However, the same study emphasized that it is important to recognize that the studies included in their meta-analysis treated PONV as a secondary outcome in their original study design, which may limit the interpretation of the results [1]. This may be a reason for the different results compared to our study. In addition, this meta-analysis [1] selected 18 studies that did not use any prophylactic antiemetic medication during anesthesia. This finding may explain and limit the result of the nonsuperiority of pregabalin compared to placebo found in our study, which used dual antiemetic therapy in all patients based on the SAMBA protocol and the Apfel scale [6]. Two other meta-analyses that evaluated the effect of pregabalin for the prevention of PONV were performed in patients undergoing hysterectomy, with conflicting results [20, 21]. It is noteworthy that all of their included studies treated PONV as a secondary outcome. Although Wang et al. [20] found a positive effect of pregabalin at controlling PONV, they emphasized that the different doses of pregabalin used and the heterogeneity of the studies were limiting factors of their meta-analysis. Ni J et al. [21] explained that they did not conclude that there was a positive effect of pregabalin in the control of PONV due to the need to obtain better-quality randomized controlled studies, as their meta-analysis was limited by the heterogeneity and size of the studies (n<100), they considered their results not clinically significant, especially in patients taking doses below 150 mg of pregabalin. Regarding adverse events related to the use of pregabalin, in our study, 19% of the patients reported sedation without other associated symptoms, but there was no significant difference between groups (P = 0.51). Regarding this side effect of sedation, meta-analyses have obtained similar results [1, 20, 21].

This study is limited by our understanding of the antiemetic mechanisms of action of pregabalin. More high-quality randomized, controlled, multicenter, double-blind clinical trials with more evaluated patients, studying the antiemetic effect of pregabalin as the primary outcome, will be needed to validate the results of the present study.

## Conclusions

The administration of 75 mg of pregabalin 2 hours before surgery and 12 hours after the first dose showed no significant difference from placebo in the reduction of episodes of PONV in patients undergoing breast reconstruction after bariatric surgery under general anesthesia. There were no significant differences between the groups in the reduction of postoperative pain, the need for antiemetic and analgesic rescue medication, the development of postoperative chronic pain, or the number of adverse events with the use of pregabalin.

## Data Availability

All relevant data are within the manuscript and its Supporting Information files.

N/A.

**Conceptualization:** Rafael Reis Fernandes, Ismar Lima Cavalcanti, Nubia Verçosa, Marcello Fonseca Salgado Filho.

**Data curatorship:** Rafael Reis Fernandes.

**Formal analysis:** Rafael Reis Fernandes, Ismar Lima Cavalcanti, Nubia Verçosa, Marcello Fonseca Salgado Filho, Rosangela Aparecida Gomes Martins.

**Investigation:** Rafael Reis Fernandes, Ismar Lima Cavalcanti, Nubia Verçosa, Marcello Fonseca Salgado Filho.

**Methodology:** Rafael Reis Fernandes, Ismar Lima Cavalcanti, Nubia Verçosa, Marcello Fonseca Salgado Filho, Rosangela Aparecida Gomes Martins, Alice Ramos Oliveira da Silva.

**Project management:** Rafael Reis Fernandes.

**Validation:** Ismar Lima Cavalcanti, Nubia Verçosa, Marcello Fonseca Salgado Filho.

**Writing – original draft:** Rafael Reis Fernandes, Ismar Lima Cavalcanti, Nubia Verçosa, Marcello Fonseca Salgado Filho.

**Writing – proofreading and editing:** Rafael Reis Fernandes, Ismar Lima Cavalcanti, Nubia Verçosa, Marcello Fonseca Salgado Filho.

